# A Novel Scoring System for Coronary Care Unit Patients: Merging SAPS 2 and Troponin I for Improved Prognostication

**DOI:** 10.1101/2025.01.15.25320621

**Authors:** Mohammad Saquib Alam, Khawaja Saifullah Zafar, Syed Hasan Amir, Shahzad F. Haque

## Abstract

**Aims:** This study aimed to develop a novel prognostic scoring system for coronary care unit (CCU) patients by integrating the Simplified Acute Physiology Score II (SAPS II) with cardiac-specific biomarkers, including Troponin I (Trop I), NT-proBNP, lactate, AST, and ALT. The objective was to enhance the prediction of in-hospital mortality by addressing limitations of existing scoring systems.

**Methods:** This prospective observational study included 25 adult patients admitted to a tertiary care hospital’s CCU with acute coronary syndrome (ACS) and acute decompensated heart failure (ADHF). Clinical and laboratory parameters, including Troponin I (Trop I), NT-proBNP, lactate, AST, ALT, and SAPS II scores, were collected upon admission. Logistic regression analysis identified independent predictors of mortality, and a new scoring system was developed. The predictive accuracy of the system was evaluated using receiver operating characteristic (ROC) analysis.

**Results:** The novel scoring system demonstrated superior discriminatory performance with an area under the ROC curve (AUC) of 0.8897 compared to SAPS II alone (AUC: 0.8493). Troponin I emerged as the most significant predictor of mortality (p < 0.05), while SAPS II showed a trend toward significance. The optimal cutoff score for the new system was determined to be 4.716, achieving a sensitivity of 75% and specificity of 94.12%. Elevated lactate, Trop I and SAPS 2 score levels were strongly associated with mortality.

**Discussion:** The new scoring system integrates systemic and cardiac-specific parameters, enhancing the predictive accuracy of in-hospital mortality in CCU patients compared to SAPS II alone. While Trop I proved highly predictive, other biomarkers (NT-proBNP, AST, ALT, lactate) did not achieve statistical significance in multivariate analysis, likely due to the limited sample size. Future validation in larger cohorts is required to confirm its generalizability and clinical utility. This study underscores the potential of combining systemic and cardiac-specific biomarkers to refine risk stratification in CCU settings, offering a robust tool for guiding clinical decision-making.

## 1. Introduction

Acute coronary syndrome (ACS), a leading cause of morbidity and mortality worldwide, often necessitates advanced care in CCUs due to its potential for severe complications such as cardiogenic shock (CS) and multiorgan dysfunction syndrome (MODS). Despite advancements in revascularization strategies and intensive care protocols, the ability to accurately predict in-hospital mortality remains suboptimal (1,2). The management of critically ill patients in coronary care units (CCUs) requires accurate and reliable prognostic tools to guide clinical decision-making and improve outcomes.

The Simplified Acute Physiology Score II (SAPS II), widely used in intensive care units (ICUs), offers robust predictive accuracy for mortality by incorporating physiological and clinical parameters used for patient in intensive care settings (3,4). However, studies suggest that SAPS II, while effective, may not fully capture the nuances of cardiac-specific pathology, especially when compared to tailored systems like the GRACE score for acute coronary syndromes (ACS). Numerous prognostic scoring systems, such as the Simplified Acute Physiology Score (SAPS III), and the Acute Physiology and Chronic Health Evaluation (APACHE) systems, have demonstrated their utility in critically ill patients, including those with AMI and CS (2–5). However, these general scoring systems often lack cardiac-specific biomarkers, potentially limiting their precision in predicting outcomes in CCU patients.

Cardiac-specific parameters, such as troponin I (Trop I) and N-terminal pro-B-type natriuretic peptide (NT-pro-BNP), have emerged as powerful predictors of adverse outcomes in acute coronary syndromes (ACS). Elevated Trop I levels are directly associated with myocardial injury, while NT-pro-BNP reflects myocardial stress and ventricular dysfunction. Additionally, other markers like lactate, aspartate aminotransferase (AST), and alanine aminotransferase (ALT) have shown potential in capturing metabolic and hepatic derangements secondary to cardiac compromise (6). Incorporating these parameters into prognostic models could enhance risk stratification and align predictions more closely with the pathophysiology of cardiac disease. However, studies suggest that SAPS II, while effective, may not fully capture the nuances of cardiac-specific pathology, especially when compared to tailored systems like the GRACE score for acute coronary syndromes (ACS) (7).

Current research highlights the limitations of general ICU scoring systems when applied to CCU patients. For instance, while SAPS II exhibits strong calibration and predictive accuracy in critically ill patients, its reliance on broad clinical parameters may lead to overestimation or underestimation of mortality in ACS-specific context. SAPS III, though more robust in predicting mortality, lacks integration of cardiac-specific biomarkers, which are critical in ACS and CS management. Moreover, the GRACE 2.0 score, specifically tailored for ACS, demonstrates high sensitivity but may falter in ICU contexts due to its limited focus on systemic organ dysfunction(7).

Given these limitations, there is a pressing need for a scoring system that merges the strengths of established prognostic models with the inclusion of cardiac-specific biomarkers. This study aims to address this gap by developing a novel scoring system for CCU patients that integrates Trop I, BNP, lactate, AST, and ALT into a SAPS II-based framework. By leveraging both systemic physiological markers and cardiac-specific parameters, we hypothesize that this new scoring system will provide improved prognostic accuracy and better guide clinical decision-making in CCU settings.

## 2. Material and Methods

### 2.1 Participants

This study included 25 patients admitted to the cardiac care unit of a tertiary care hospital in North India, presenting with acute coronary syndrome (ACS) and acute decompensated heart failure (ADHF). All patients were adults aged ≥18 years. Patients not meeting these criteria or presenting with other conditions were excluded from the study.

### 2.2 Material

Clinical evaluations and investigations were conducted for all included patients at the time of admission. The parameters assessed included:

- **Vital signs:** pulse rate and systolic blood pressure.
- **Laboratory investigations:**
  ∘Complete blood count (CBC)
  ∘Blood gas analysis
  ∘Renal function tests (RFT)
  ∘Serum electrolytes
  ∘Liver function tests (LFT)
  ∘Troponin I (Trop I)
  ∘N-terminal pro B-type natriuretic peptide (NT-proBNP)
  ∘Serum lactate levels
- **Severity assessment:** Simplified Acute Physiology Score II (SAPS II) was calculated for all patients upon admission.

The final clinical outcomes of the patients were recorded as either “improved” or “expired” during the course of their hospital stay.

### 2.3 Procedure

The study was a hospital based prospective observational study. Patients underwent a detailed clinical evaluation upon admission, including measurement of vital parameters such as pulse rate and systolic blood pressure. Blood samples were collected for laboratory investigations, which were analyzed as part of the diagnostic and prognostic assessment. The SAPS II score was calculated for each patient to determine the severity of illness and predict hospital mortality. Clinical follow-up was performed throughout the hospital admission to record the final outcomes (Fig.1).

**Fig 1:**
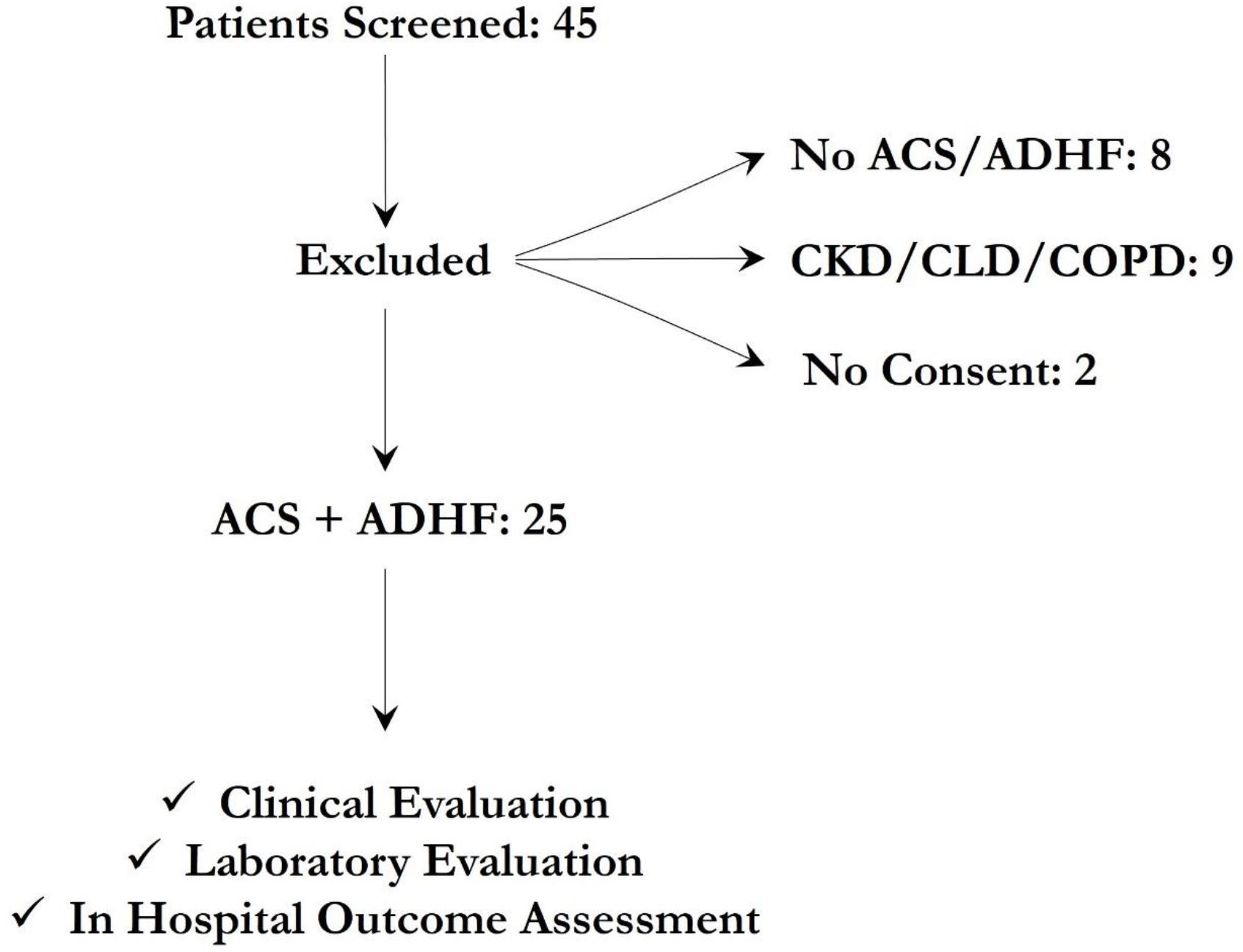
This flowchart illustrates the study design for patients admitted to the cardiac care unit (CCU) in a tertiary care hospital in North India. Patients with acute coronary syndrome (ACS) and acute decompensated heart failure (ADHF) aged ≥18 years were included. Exclusion criteria encompassed non-ACS/ADHF conditions, age <18 years, severe comorbidities (chronic kidney disease (CKD), Chronic Liver Disease (CLD) and Chronic Obstructive Pulmonary Disease), and unwilling patients. After applying the criteria, 25 patients formed the final study population. Outcomes were assessed as “improved” or “expired” during hospital admission.

### 2.4 Statistical Analysis

Statistical analysis was performed to evaluate the association between clinical parameters, SAPS II scores, and patient outcomes. Descriptive statistics were used to summarize the demographic and clinical characteristics of the study population, with continuous variables expressed as mean ± standard deviation (SD) and categorical variables presented as proportions. Univariate analysis was conducted using chi-square tests for categorical variables and t-tests or Mann-Whitney U tests for continuous variables to identify significant differences between outcome groups. Logistic regression analysis was employed to identify independent predictors of mortality and to develop a predictive scoring system. The accuracy and discriminatory power of the scoring system were evaluated using receiver operating characteristic (ROC) analysis, with the area under the curve (AUC) calculated to assess its ability to differentiate between patients who improved and those who expired. A p-value of<0.05 was considered statistically significant for all analyses.

## 3. Results

The study conducted a detail analysis of the SAPS score, cardiac biomarkers and other laboratory parameters (Lactate, NT pro BNP, AST and ALT) of 25 patients included in the study and followed them during the period of hospitalization to look for in hospital outcome.

### 3.1 Demographics

A total of 25 patients admitted to the cardiac care unit (CCU) with acute coronary syndrome (ACS) and acute decompensated heart failure (ADHF) were included in the study. The mean age of the patients was 62 ± 14.1 years, with 48% being male. The baseline clinical and laboratory parameters are summarized in Table 1. Key findings included a mean systolic blood pressure of 95.6 ± 37.6 mmHg, mean serum lactate levels of 5.5± 4.3 mmol/L, and mean SAPS II score of 41.5 ± 12.7.

**Table 1:** Baseline Characteristics.

| Parameter | Mean ± SD |
| --- | --- |
| Age (yrs) | $62.36 \pm 14.09$ |
| Heart Rate (bpm) | $92.20 \pm 31.65$ |
| Systolic Blood Pressure (mmHg) | $95.60 \pm 37.64$ |
| Temperature ($^{\circ}$F) | $98.58 \pm 0.06633$ |
| Blood Urea Nitrogen (mg/dL) | $32.44 \pm 21.99$ |
| Urine Output (mL/24 hrs.) | $629.2 \pm 417.8$ |
| Sodium (mEq/L) | 137.6 ± 5.438 |
| Potassium (mEq/L) | 4.320 ± 0.9522 |
| Bicarbonate Ion (mEq/L) | 18.12 ± 6.011 |
| Bilirubin (mg/dL) | 1.500 ± 0.9101 |
| Total Leukocyte Count (10<sup>3</sup>/uL) | 12.63 ± 5.015 |
| Troponin I (ng/L) | 7984 ± 12623 |
| NT pro BNP ( | 15037 ± 9296 |
| Aspartate Aminotransferase (U/L) | 1457 ± 2440 |
| Alanine Aminotransferase (U/L) | 964.2 ± 1758 |
| Creatinine (mg/dL) | 2.392 ± 1.686 |
| Lactate (mEq/L) | 5.512 ± 4.305 |
| SAPS Score | 41.52 ± 12.65 |

### 3.2 Comparison of Parameters between Survivors and Non-Survivors

In the analysis of clinical and biochemical parameters, several variables were found to be statistically significant predictors of mortality (p < 0.05). Non-survivors exhibited significantly lower heart rates compared to survivors (70.9 ± 36.7 bpm vs. 102.2 ± 24.1 bpm, p = 0.0275), suggesting an association between bradycardia and adverse outcomes (Fig 2A). Lactate levels were markedly elevated in non-survivors (8.3 ± 4.8 mEq/L) compared to survivors (4.2 ± 3.4 mEq/L, p = 0.0375), reflecting greater metabolic derangement in those who expired (Fig 2B). Similarly, bicarbonate levels were significantly lower among non-survivors (14.1 ± 6.2 mEq/L vs. 20.0 ± 5.1 mEq/L, p = 0.0325), indicative of a higher degree of acidosis in this group (Fig 2C). Troponin I levels, a marker of myocardial injury, were substantially elevated in non-survivors (19895 ± 16885 ng/L vs. 2379 ± 3156 ng/L, p = 0.0072), correlating with worse cardiac outcomes (Fig 2D). Furthermore, the SAPS II score, a measure of illness severity, was significantly higher in non-survivors (50.3 ± 9.1) than in survivors (37.4 ± 12.2, p = 0.0041), underscoring its prognostic value in this cohort (Fig 2E) (Table 2). These findings highlight the critical role of these variables in predicting mortality and assessing the severity of illness in patients with acute coronary syndrome and acute decompensated heart failure.

**Table 2:** Comparison of Clinical and Laboratory Parameters between survivors and non survivors.

| Parameters | Survivors | Non-Survivors | P value |
| --- | --- | --- | --- |
| Age (yrs) | 60.1 ± 14.8 | 67.1 ± 11 | 0.4804 |
| Heart Rate (bpm) | 102.2 ± 24.1 | 70.9 ± 36.7 | **0.0275** |
| Systolic Blood Pressure (mmHg) | 87.4 ± 19.0 | 113 ± 59.4 | 0.4457 |
| Lactate (mEq/L) | 4.2 ± 3.4 | 8.3 ± 4.8 | **0.0375** |
| Bicarbonate Ion (mEq/L) | 20.0 ± 5.1 | 14.1 ± 6.2 | **0.0325** |
| Troponin I (ng/L) | 2379 ± 3156 | 19895 ± 16885 | **0.0072** |
| NT pro BNP (pg/mL) | 15065 ± 9012 | 14979 ± 10521 | 0.8967 |
| Aspartate Aminotransferase (U/L) | 1406 ± 2432 | 1564 ± 2622 | 0.0678 |
| Alanine Aminotransferase (U/L) | 924 ± 1676 | 1050 ± 2039 | 0.3823 |
| SAPS 2 score | 37.4 ± 12.2 | 50.3 ± 9.1 | **0.0041** |

**Table 3:** Performance Metrics of the Predictive Scoring System at Various Cutoff Scores.

| Cut off Score | Sensitivity | 95% CI | Specificity | 95% CI | Likelihood Ration |
| --- | --- | --- | --- | --- | --- |
| > 3.579 | 75.00 | 40.93% to 95.56% | 76.47 | 52.74% to 90.44% | 3.188 |
| > 3.928 | 75.00 | 40.93% to 95.56% | 82.35 | 58.97% to 93.81% | 4.250 |
| > 4.203 | 75.00 | 40.93% to 95.56% | 88.24 | 65.66% to 97.91% | 6.375 |
| > 4.716 | 75.00 | 40.93% to 95.56% | 94.12 | 73.02% to 99.70% | 12.75 |
| > 5.314 | 62.50 | 30.57% to 86.32% | 94.12 | 73.02% to 99.70% | 10.63 |

**Table 4:** Diagnostic Performance Metrics for the Cutoff Score of 4.716.

| Metric | Value | 95% Confidence Interval |
| --- | --- | --- |
| Sensitivity | 75.00% | 40.93% to 95.56% |
| Specificity | 94.12% | 73.02% to 99.70% |
| Positive Predictive Value (PPV) | 85.71% | 48.69% to 99.27% |
| Negative Predictive Value (NPV) | 88.89% | 67.20% to 98.03% |

**Fig 2:**
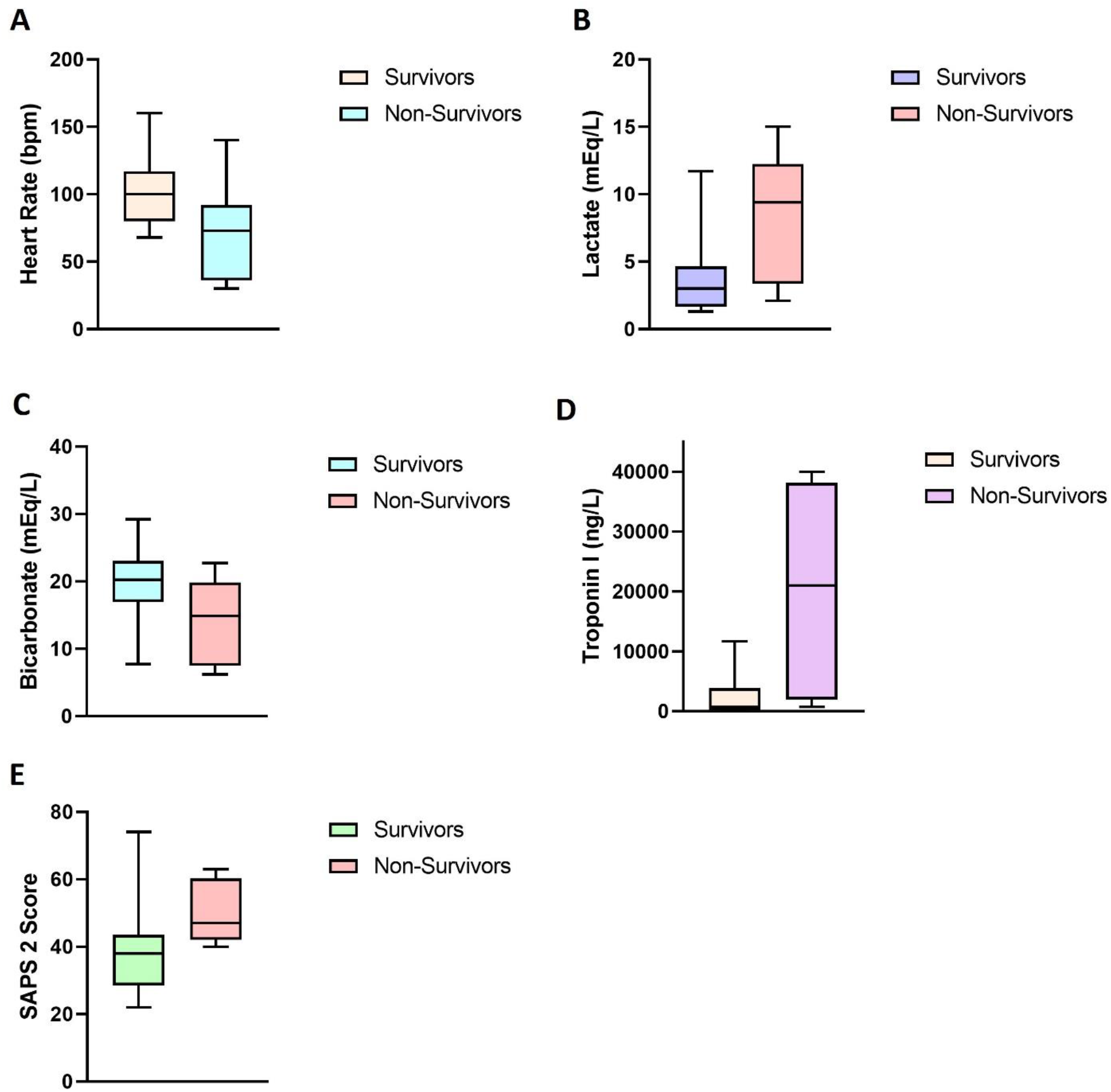
Illustrates the comparison of key clinical and biochemical parameters between survivors and non-survivors in the study population, presented as box-and-whisker plots. Survivors demonstrated a significantly higher heart rate (Panel A) compared to non-survivors. Lactate levels (Panel B) were markedly elevated in non-survivors. Similarly, bicarbonate levels (Panel C) were significantly lower in non-survivors. Troponin I levels (Panel D) were substantially higher in non-survivors. SAPS II scores (Panel E) were significantly higher among non-survivors, underscoring their utility as a prognostic tool in predicting mortality.

**Fig 3:**
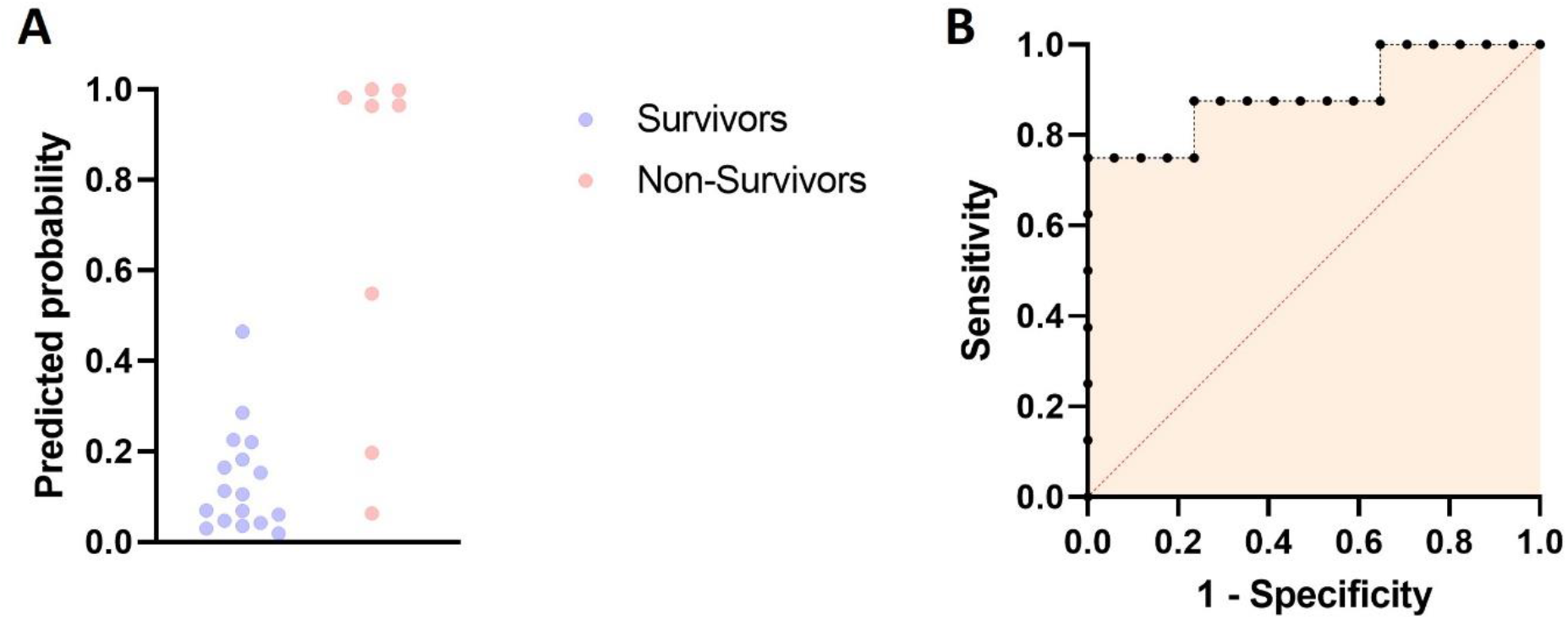
The figure consists of two panels visualizing the predictive performance of a logistic regression model for mortality outcomes. Panel A shows a scatterplot of predicted probabilities for survivors (blue dots) and non-survivors (red dots). Survivors predominantly cluster around lower predicted probabilities, whereas non-survivors are concentrated near higher probabilities, indicating the model’s ability to distinguish between the two groups. Panel B displays the Receiver Operating Characteristic (ROC) curve, illustrating the trade-off between sensitivity and 1-specificity across varying cutoff points. The curve’s high placement suggests strong discriminatory power, with an area under the curve (AUC) close to 1, indicative of excellent model performance.

**Fig 4:**
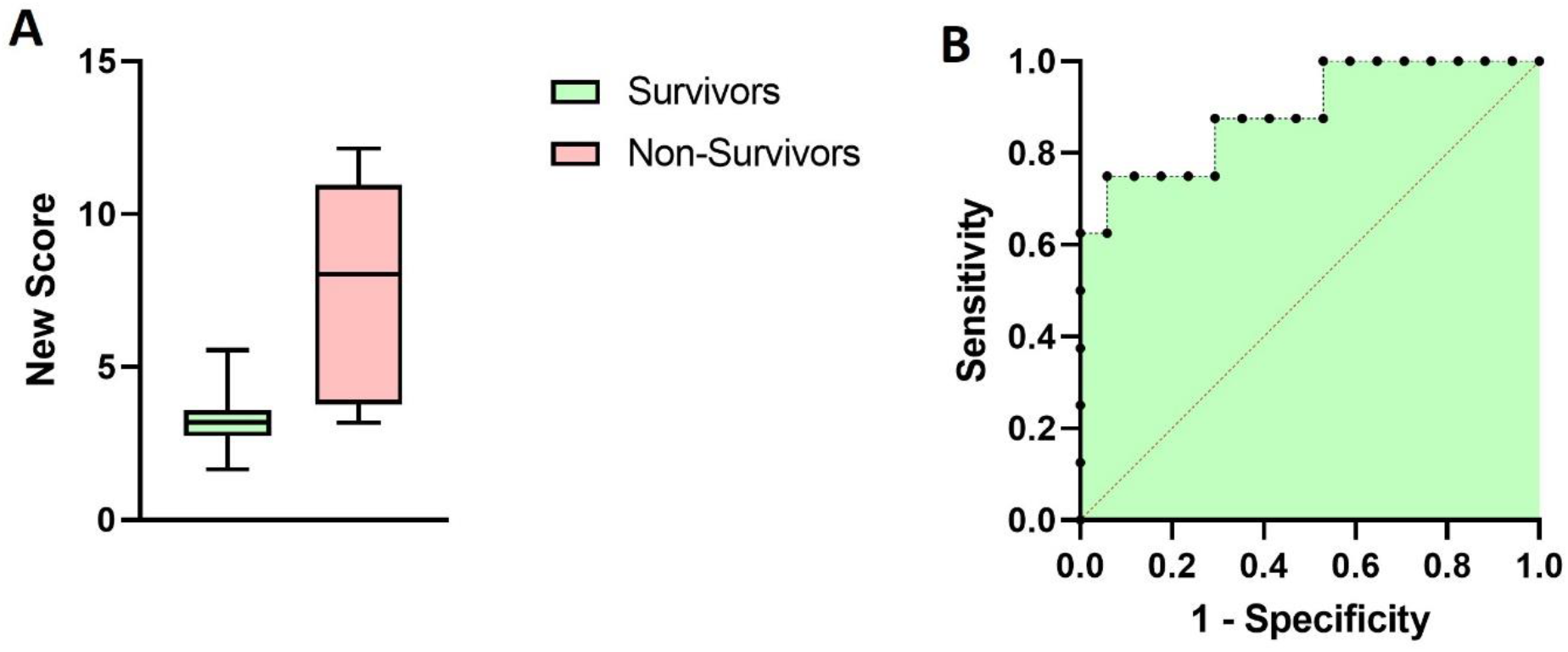
Illustrates the performance of the newly developed scoring system in predicting outcomes. **Panel A** shows a box-and-whisker plot comparing the scores between survivors (green) and non-survivors (pink), with significantly higher scores in non-survivors, indicating strong discriminative ability. **Panel B** displays the ROC curve, demonstrating the trade-off between sensitivity and specificity across different cutoff points.

**Fig 5:**
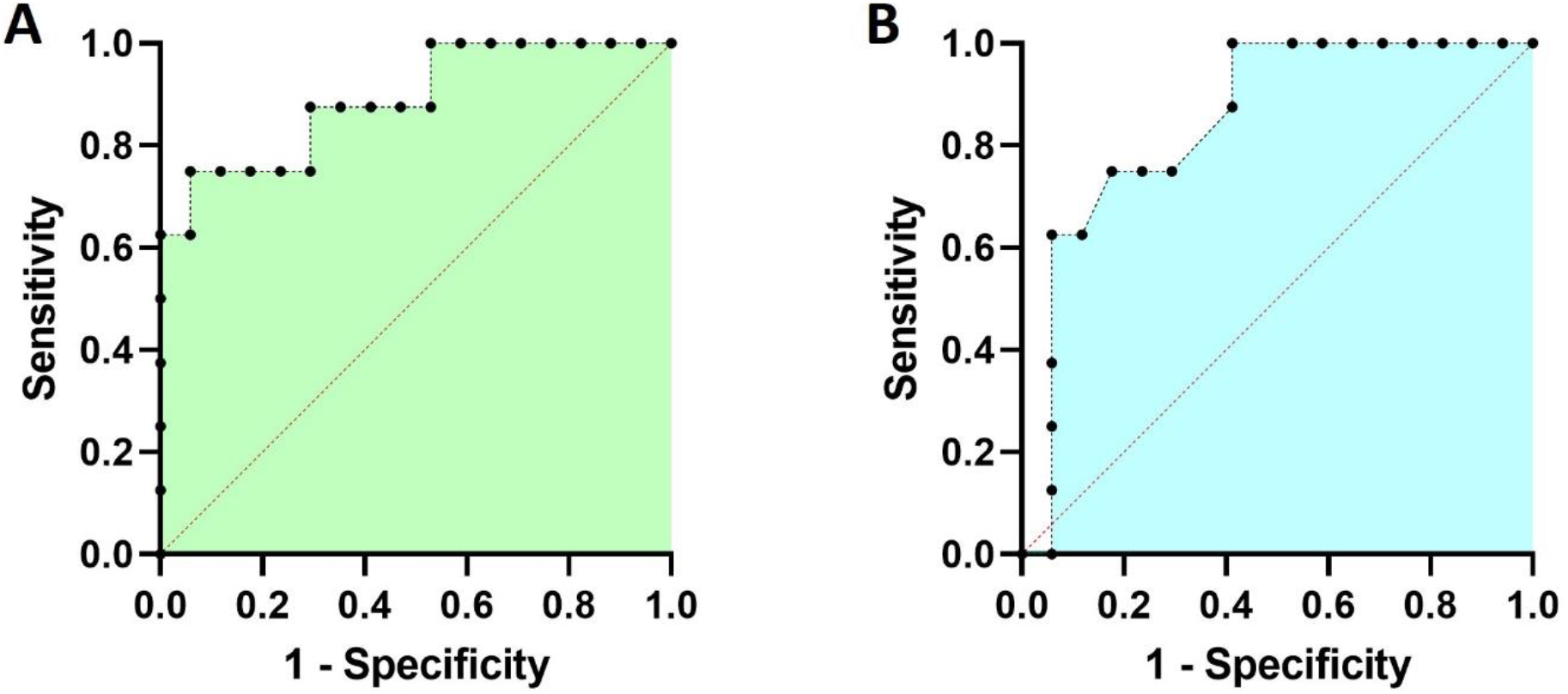
This figure illustrates the Receiver Operating Characteristic (ROC) curves for the predictive performance of the newly developed scoring system (**Panel A**) and the SAPS 2 score (**Panel B**) in distinguishing survivors from non-survivors. In **Panel A**, the ROC curve for the new scoring system demonstrates excellent discriminative ability, with a high area under the curve (AUC) of 0.8897, reflecting strong predictive accuracy. In **Panel B**, the ROC curve for the SAPS 2 score shows good performance, with an AUC of 0.8493. While both models exhibit strong discriminatory power, the new scoring system outperforms the SAPS 2 score in terms of AUC and overall separation between sensitivity and 1-specificity, suggesting its superior utility for predicting mortality outcomes.

### 3.3 Multiple Logistic Regression Analysis

A multiple logistic regression model was developed to predict mortality outcomes using six clinical parameters: Trop I, NT-proBNP, AST, ALT, lactate, and SAPS 2 score. The binary outcome variable was mortality, with 0 indicating survival and 1 indicating death. The model demonstrated excellent discriminatory performance, with an area under the receiver operating characteristic (ROC) curve (AUC) of 0.8897 (95% CI: 0.7290 to 1.000, p = 0.002), indicating a strong ability to distinguish between survivors and non-survivors. Troponin I emerged as the most significant predictor of mortality (β=0.000196, p<0.05), with higher levels associated with an increased likelihood of death. Although the SAPS 2 score showed a trend toward significance (β=0.07413, p=0.08), it did not reach statistical significance; however, higher scores were associated with a potential increase in mortality risk. The remaining predictors, including NT-proBNP, AST, ALT, and lactate, were not statistically significant contributors to the model (p > 0.05). These results highlight the importance of Trop I as a key biomarker in predicting mortality, while the SAPS 2 score may also contribute meaningfully to risk stratification but did not display statistical significance. The regression model demonstrated good discriminatory power, with an area under the receiver operating characteristic (ROC) curve (AUC) of 0.8897 (95% CI: 0.7290 to 1.000, p = 0.002). This indicates excellent performance in distinguishing between survivors and non-survivors.

### 3.4 New Scoring System Formation

A novel predictive scoring system was developed based on the results of the multiple logistic regression analysis. Significant clinical parameters, including Trop I and the SAPS 2 score, were identified as the strongest predictors of mortality, as both demonstrated meaningful associations with the outcome. Troponin I had a regression coefficient (β) of 0.000196 (p<0.05), indicating that higher levels were associated with increased mortality risk. Similarly, the SAPS 2 score had a coefficient of 0.07413, with a trend toward significance (p=0.08), suggesting that higher scores corresponded to worse outcomes. Other variables, such as NT-proBNP, AST, ALT, and lactate, were not statistically significant in the regression model and were excluded from the scoring system. To create an interpretable and clinically usable scoring system, the coefficients of Trop I and the SAPS 2 score were rescaled and rounded to whole numbers for simplicity. The scoring formula was derived as follows:

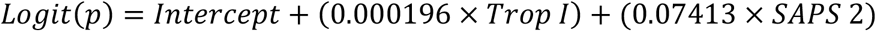

This formula assigns weight to each predictor proportional to its impact on mortality, with Trop I contributing 0.000196 points per unit and the SAPS 2 score contributing 0.07413 points per unit. The intercept from the logistic regression equation was adjusted to ensure that the minimum possible score starts at 0, improving interpretability.

Based on the analysis of sensitivity, specificity, and likelihood ratios, the optimal cutoff for the newly developed scoring system was determined to be 4.716. At this threshold, the scoring system achieves a sensitivity of 75.0**%** (95% CI: 40.93–95.56%) and a specificity of 94.12% (95% CI: 73.02–99.70%). The positive likelihood ratio (LR+) at this cutoff is 12.75, indicating excellent discriminatory power. This threshold balances the ability to correctly identify patients at risk of mortality (sensitivity) while minimizing false positives (high specificity). Selecting this cutoff ensures that the majority of patients at high risk of mortality are identified, while maintaining a low rate of false-positive classifications, which is crucial in clinical contexts where unnecessary interventions carry significant risks or costs.

### 3.5 New Score Validation

The cutoff score of 4.716 for the new scoring system was validated using chi-square analysis. The test revealed a significant association between the score and mortality (χ2=12.89, p=0.0003), confirming the predictive utility of this cutoff. The odds ratio (OR) for mortality at scores >4.716 was 48.00 (95% CI: 4.368–567.7), indicating that patients with scores above the threshold had a significantly higher likelihood of mortality. The sensitivity and specificity of the cutoff were 75.0% (95% CI: 40.93– 95.56%) and 94.12% (95% CI: 73.02–99.70%), respectively, with a likelihood ratio of 12.75, demonstrating strong discriminative power. The positive predictive value (PPV) was 85.71%, while the negative predictive value (NPV) was 88.89%, supporting the ability of the score to accurately classify patients into high- and low-risk categories. The analysis emphasized on the clinical relevance of this scoring system for identifying patients at risk of mortality. These results highlight the robust predictive performance and clinical utility of the cutoff score.

### 3.6 Comparison with SAPS-2 Score

Based on the analysis of sensitivity, specificity, and likelihood ratios, the optimal cutoff for the SAPS 2 score was selected as 44.5. At this threshold, the sensitivity was 75.0% (95% CI: 40.93%–95.56%) and the specificity was 82.35% (95% CI: 58.97%–93.81%), with a positive likelihood ratio (LR+) of 4.250. This cutoff offers a balanced approach, providing robust sensitivity for identifying patients at risk of mortality while maintaining high specificity to minimize false positives.

When comparing the SAPS 2 score to the newly developed scoring system, the new score demonstrated superior performance. At its optimal cutoff of 4.716, the new score achieved a similar sensitivity of 75.0% (95% CI: 40.93%–95.56%) but a higher specificity of 94.12% (95% CI: 73.02%– 99.70%) and an excellent LR+ of 12.75, compared to the SAPS 2 score’s LR+ of 4.250. Furthermore, the new scoring system had a higher area under the curve (AUC) of 0.8897 compared to 0.8493 for the SAPS 2 score, highlighting its superior ability to discriminate between survivors and non-survivors. These findings suggest that the new scoring system provides better predictive accuracy and clinical utility than the SAPS 2 score in patients of ACS with ADHF. Validation in larger and independent datasets is warranted to confirm its generalizability and effectiveness in diverse patient populations.

## 4. Discussions

This study highlights the development of a novel scoring system that integrates cardiac-specific biomarkers with the Simplified Acute Physiology Score II (SAPS II) to enhance prognostic accuracy in coronary care unit (CCU) patients. By incorporating Troponin I (Trop I), NT-proBNP, lactate, AST, and ALT, this system provides a more comprehensive evaluation of cardiac and systemic health, surpassing the predictive capabilities of SAPS II alone.

Previous research has underscored the utility of SAPS II in critically ill patients, particularly in intensive care unit (ICU) setting (2). However, its limitations in capturing cardiac-specific parameters have been noted. For instance, Zheng et al. demonstrated the superior predictive value of SAPS III in coronary care units, attributed to its advanced multinational database and inclusion of more nuanced clinical variables(3). Similarly, the GRACE 2.0 score has shown high sensitivity for acute coronary syndromes but lacks systemic physiological parameters critical in ICU setting (7).

In line with the findings of Silva-Obregon et. Al where SAPS II outperformed APACHE II and GRACE 2.0 in terms mortality prediction in ST-segment elevation myocardial infarction (STEMI) patients (7), our study confirms SAPS II’s foundational strength but enhances it by adding markers reflective of cardiac stress and metabolic dysfunction. By integrating Trop I and NT-proBNP, biomarkers strongly associated with adverse cardiac outcomes, and lactate, which reflects systemic hypoperfusion and metabolic derangement, we like to develop the novel score to addresses gaps identified in existing models.

In our cohort, Trop I emerged as the most significant predictor of mortality, consistent with its established role as a marker of myocardial injury(8). Elevated lactate levels were also strongly associated with poor outcomes, underscoring its relevance as a marker of critical illness. These findings align with Buerke et al., who emphasized metabolic derangements in AMI-related complications like cardiogenic shock (1).

However, after performing multivariate logistic regression only Trop I came out as a predictor of mortality in out cohort, while NT-proBNP, Lactate, AST, and ALT were included in the initial model, their contributions to mortality prediction were not statistically significant in multivariate analysis. This contrasts with findings by Jasiewicz et al., who highlighted elevated transaminases as indicators of systemic and hepatic stress in acute coronary syndromes(6). These discrepancies may stem from our smaller sample size or differences in population characteristics.

The novel scoring system including SAPS 2 score combined with Trop I demonstrated superior discriminatory power (AUC: 0.8897) compared to SAPS II alone (AUC: 0.8493). Its optimal cutoff score (4.716) achieved a sensitivity of 75% and specificity of 94.12%, outperforming SAPS II in identifying high-risk patients. These findings suggest that integrating cardiac biomarkers into established scoring systems can significantly enhance prognostic accuracy, a notion supported by Kellner et al., who advocated for tailored scoring systems in cardiogenic shock (2).

While promising, the study is limited by its small sample size and single-center design, which may affect generalizability. Furthermore, the exclusion of dynamic changes in biomarkers and scores during hospitalization restricts the evaluation of their temporal predictive value. Future studies should validate this scoring system in larger, multicenter cohorts and explore its application in diverse cardiac populations.

This study reinforces the need for tailored scoring systems that combine systemic and cardiac-specific parameters. The proposed scoring system builds on the strengths of SAPS II while addressing its limitations, offering a robust tool for risk stratification in CCU patients. Its superior performance highlights the potential for integrating biomarkers like Trop I into predictive models, paving the way for more precise and clinically actionable prognostication in acute cardiac care.

## Data Availability

All data produced in the present study are available upon reasonable request to the authors

